# Serum Tumor Marker Profiles in Interstitial Lung Diseases: Implications for Differential Diagnosis and Disease Severity Assessment

**DOI:** 10.64898/2026.01.13.26344081

**Authors:** Yingying Du, Xuxiang Song, Qunli Ding

## Abstract

**Background:** Interstitial lung diseases (ILDs), including idiopathic pulmonary fibrosis (IPF) and connective tissue disease-associated ILD (CTD-ILD), share similar features that complicate diagnosis. Tumor markers are often elevated in ILD, yet their diagnostic utility remains unclear.

**Methods:** This retrospective study included ILD patients hospitalized between 2018 and 2025. Serum levels of alpha-fetoprotein, carcinoembryonic antigen (CEA), carbohydrate antigen (CA) 199, CA125, CA153, neuron-specific enolase (NSE), and cytokeratin 19 fragment (CYFRA 21-1) were analyzed. Arterial blood gases and erythrocyte sedimentation rates (ESRs) were also collected. Statistical analyses involved the Kruskal–Wallis test, Dunn’s post hoc test, Spearman’s correlation, logistic regression, and receiver operating characteristic (ROC) curve analysis.

**Results:** CEA, CA199, and CA125 levels varied significantly among ILD subtypes (all *p* < 0.05). NSE differed among CTD-ILD subgroups (*p* = 0.0409). In IPF, CEA and NSE correlated inversely with PaO_2_ (r = −0.1556, *p* = 0.0380; r = −0.2205, *p* = 0.0031). In CTD-ILD, NSE correlated negatively with PaCO_2_ (r = −0.1811, *p* = 0.016), and CYFRA 21-1 with PaO_2_ (r = −0.1999, *p* = 0.0078). A diagnostic model incorporating CEA, CA199, sex, age, smoking, PaO_2_, and ESR differentiated IPF from CTD-ILD with an AUC of 0.833 (95% CI: 0.790–0.876), showing 73.6% sensitivity and 82.4% specificity at a cutoff of 0.569, outperforming single markers.

**Conclusion:** CEA, CA199, and CA125 aid in distinguishing ILD subtypes, while CEA, NSE, and CYFRA 21-1 correlate with impaired gas exchange. The combined clinical and biomarker model demonstrated superior performance in discriminating IPF from CTD-ILD, highlighting its clinical potential.

## 1. Introduction

A diverse collection of diffuse parenchymal lung disorders known as interstitial lung diseases (ILDs) are distinguished by differing degrees of fibrosis and inflammation, which eventually lead to progressive respiratory failure^[1]^. Although ILDs have different causes, many patients present similar symptoms, such as chronic cough and shortness of breath during exercise. Idiopathic pulmonary fibrosis (IPF) is pathologically defined by the usual interstitial pneumonia (UIP) pattern and typically follows a relentlessly progressive course despite therapy, for which anti-fibrotic agents are the mainstay of treatment^[2]^. In contrast, connective tissue disease-related ILDs (CTD-ILDs) occur in the context of autoimmune disorders such as systemic sclerosis (SSc), rheumatoid arthritis (RA), and systemic lupus erythematosus (SLE)^[3]^. CTD-ILDs are typically associated with the nonspecific interstitial pneumonia (NSIP) pattern and may respond to immunosuppressive therapy^[4]^. Because of these divergent therapeutic approaches, accurate differentiation between IPF and CTD-ILD is of great clinical importance. However, some patients may present with interstitial lung disease as the initial or even sole manifestation in the early stage of connective tissue disease, during which serological antibody tests can yield negative or inconclusive results^[5]^. Such seronegative or atypical presentations often delay diagnosis and make it challenging to distinguish CTD-ILD from IPF. Moreover, a subset of CTD-ILDs can exhibit a UIP pattern that is radiologically and histopathologically indistinguishable from IPF, further complicating diagnostic accuracy and often being associated with poorer prognoses. Given this diagnostic uncertainty and its direct therapeutic implications, there is an urgent need for objective and reliable biomarkers that can assist in differentiating ILD subtypes and guide appropriate treatment selection. Accordingly, recent research has focused on tumor markers and other serum indicators as potential tools for improving diagnostic precision and evaluating disease severity ^[6]^.

Tumor markers refer to substances abnormally produced by malignant tumor cells or substances generated by the host in response to tumor stimulation, which can reflect the occurrence and development of tumors and be used to monitor the tumor’s response to treatment^[7]^. In clinical practice, markers such as cytokeratin fragment 21-1 (CYFRA21-1), carcinoembryonic antigen (CEA), and cancer antigen (CA) 125 are commonly used to help diagnose and treat various cancers, including those of the lung, gastrointestinal tract, and ovaries^[8]^.

An increasing number of studies have shown that tumor markers can be expressed not only in cancers but also in noncancer conditions, such as long-term inflammation, autoimmune diseases, and lung fibrosis^[9, 10]^. Some reports have shown that people with ILD may have high tumor marker levels even when no cancer is present^[11–14]^. Beyond general inflammatory responses, the elevation of these markers in ILD may reflect disease-specific pathological mechanisms. For example, CYFRA 21-1, is released during alveolar epithelial cell injury and apoptosis, processes that are prominent in ILD^[15]^. CA125, which is secreted by mesothelial and epithelial cells, has been reported to increase in response to fibroblast activation and epithelial–mesenchymal transition (EMT), both of which contribute to extracellular matrix deposition and fibrosis progression^[16]^. Therefore, the aim of this study was to evaluate the combined utility of a panel of routine and easily accessible tumor markers for differentiating IPF from CTD-ILD and for assessing disease severity. Compared with more expensive markers such as KL-6, these routine tumor markers are widely available and cost-effective in clinical laboratories, and may be particularly valuable in ILD patients with negative autoimmune serology or atypical clinical features.

## 2. Methods

### 2.1. Subjects

This study included patients with different types of ILD who were admitted to the First Affiliated Hospital of Ningbo University from January 1, 2018 to July 1, 2025. The study followed the principles set forth in the Declaration of Helsinki and was approved by the Institutional Review Board of the First Hospital Affiliated with Ningbo University. (Approval No.2025 Research No. 129RS). The clinical data used in this retrospective study were accessed from the electronic medical record system of The First Affiliated Hospital of Ningbo University on July 15, 2025. All data were de-identified prior to analysis; the authors did not have access to any information that could identify individual participants. Figure 1 shows the flowchart of the study.

**Figure 1.**
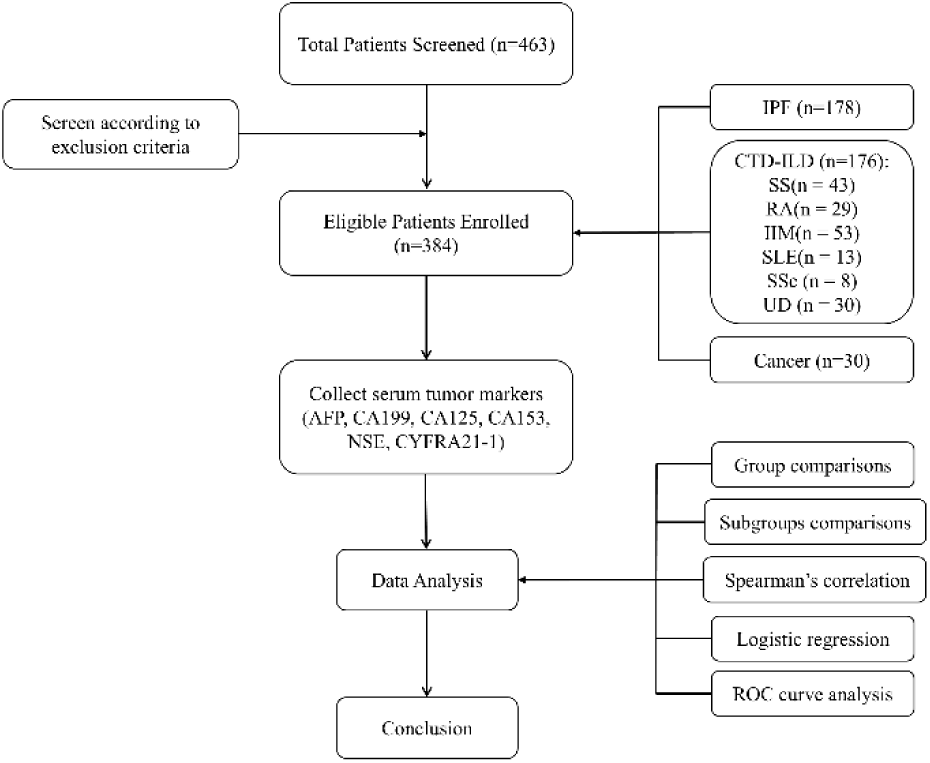
Flowchart of the study.

IPF was diagnosed according to the 2018 ATS/ERS/JRS/ALAT clinical practice guideline ^[2]^, based on compatible clinical manifestations and HRCT findings consistent with a UIP pattern. Patients in the cancer group were those diagnosed with interstitial lung disease (ILD) who had a concurrent malignant tumor, confirmed by histopathological. CTD was diagnosed in patients who fulfilled internationally recognized classification criteria for specific connective tissue diseases, including rheumatoid arthritis (RA; 2010 ACR/EULAR criteria)^[17]^, primary Sjögren’s syndrome (SS; 2002 revised international classification criteria)^[18]^, idiopathic inflammatory myopathy (IIM; 2004 ENMC criteria)^[19]^, systemic lupus erythematosus (SLE; 2019 EULAR/ACR classification criteria)^[20]^, and systemic sclerosis (SSc; 2013 ACR/EULAR classification criteria)^[21]^. Patients with undifferentiated CTD (UD) who had clinical features suggestive of CTD but did not meet criteria for a specific disease were also included. CTD-ILD was defined by: (1) a confirmed diagnosis of CTD; (2) respiratory symptoms such as cough, dyspnea, or Velcro crackles; (3) HRCT findings indicative of ILD, such as subpleural honeycombing or ground-glass opacities; (4) pulmonary function tests showing restrictive ventilation and/or impaired diffusion; and (5) exclusion of other causes including drug-induced, neoplastic, or exposure-related ILD. The cancer group comprised patients with interstitial lung disease who had a concurrent diagnosis of malignancy confirmed by clinical, imaging, or pathological evidence.

The exclusion criteria were as follows: chronic inflammatory diseases such as cirrhosis, COPD, chronic kidney disease, or other diseases known to affect tumor marker levels; active tuberculosis or other severe respiratory infections; and severe dysfunction of major organs. Patients in the IPF and CTD-ILD groups with a history of malignancy or active cancer were excluded.

### 2.2. Collection of Laboratory Data

The clinical data collected included age, sex, and smoking history. Laboratory data were retrieved from the electronic medical records of all enrolled patients. Serum tumor marker levels—including alpha-fetoprotein (AFP), CEA, CA199, CA125, CA153, neuron-specific enolase (NSE), and CYFRA21-1—were measured at the time of hospital admission, prior to the initiation of any disease-specific treatments such as immunosuppressive or antifibrotic therapy. The normal values for these tumor markers were set as follows: AFP <7 ng/mL, CEA <5.00 ng/mL, CA199 <25 U/mL, CA125 <23 U/mL, CA153 <15 U/mL, NSE <16.5 ng/mL and CYFRA21-1 <3.3 ng/mL. In addition, arterial blood gas parameters, including PaO_2_ and PaCO_2_, as well as the ESR, were collected within 24 hours of admission. All laboratory tests were performed in the central clinical laboratory of The First Affiliated Hospital of Ningbo University via standardized procedures. Data quality was independently verified by two investigators to ensure accuracy and completeness.

### 2.3. Statistical analysis

All the statistical analyses were performed via R software (version 4.5.0). The Shapiro–Wilk test was used to assess the normality of continuous variables. Variables following a normal distribution are presented as mean ± standard deviation (SD), whereas non-normally distributed variables are presented as median with interquartile range (IQR). Missing data were addressed using multiple imputation by chained equations with 5 imputed datasets. Analyses were performed on each dataset and results were pooled according to Rubin’s rules. For comparisons among more than two groups, the Kruskal–Wallis test was applied. When significant, pairwise comparisons were performed via Dunn’s test with Benjamini‒Hochberg adjustment to control for multiple testing. For categorical variables, differences among groups were assessed via the chi-square (χ²) test. A two-sided *p* value < 0.05 was considered statistically significant. Correlations between tumor markers and clinical parameters, including arterial blood gas values (PaO_2_ and PaCO_2_) and the ESR, were assessed via Spearman’s rank correlation coefficient. To identify tumor markers significantly associated with the likelihood of IPF versus CTD-ILD, univariate logistic regression analysis was conducted. Markers that reached statistical significance were further included in a multivariate logistic regression model to identify independent predictors. The results are presented as odds ratios (ORs) with 95% confidence intervals (*CIs*). The diagnostic performance of each tumor marker in differentiating between IPF and CTD-ILD patients was evaluated via receiver operating characteristic (ROC) curve analysis.

## 3. Results

A total of 384 patients were analyzed, including 178 patients with IPF, 176 patients with CTD-ILD, and 30 patients with ILD related to cancer. The CTD-ILD group included subtypes such as Sjögren’s syndrome (n = 43), rheumatoid arthritis (n = 29), idiopathic inflammatory myopathies (n = 53), systemic lupus erythematosus (n = 13), systemic sclerosis (n = 8), and undifferentiated connective tissue disease (n = 30).

### 3.1. Demographic characteristics of the study participants and results of group comparisons

A total of 384 patients were included in the study, consisting of 178 patients with IPF, 176 with CTD-ILD, and 30 with cancer-associated ILD. The median age differed significantly among the three groups (Kruskal‒Wallis test, χ² = 56.324, df = 2, *p*< 0.001), with the cancer group having the highest median age (74 years, IQR: 69.5–81.8), followed by the IPF group (70 years, IQR: 63–76) and the CTD-ILD group (62.5 years, IQR: 54–70). The sex distribution varied significantly across groups (Pearson’s chi-square test, χ² = 83.513, df = 2, *p* < 0.001), with a greater proportion of males in the IPF and cancer groups than in the predominantly female CTD-ILD group. Additionally, smoking history differed significantly among the groups (χ² = 48.14, df = 2, *p* < 0.001), with the IPF and cancer groups having a greater prevalence of smokers than the CTD-ILD group.

Tumor marker analysis revealed that AFP, CA153, NSE, and CYFRA21.1 did not differ significantly among groups (all p > 0.05). In contrast, CEA, CA199, and CA125 showed significant differences. Dunn’s multiple comparisons with Benjamini–Hochberg correction indicated: CEA was higher in the cancer vs CTD-ILD group and in CTD-ILD vs IPF; CA199 was higher in CTD-ILD vs IPF, while the difference between cancer and CTD-ILD did not reach statistical significance (adjusted p = 0.052); CA125 differed between cancer vs CTD-ILD and CTD-ILD vs IPF. The results are shown in Table1 and Figure 2.

**Figure 2.**
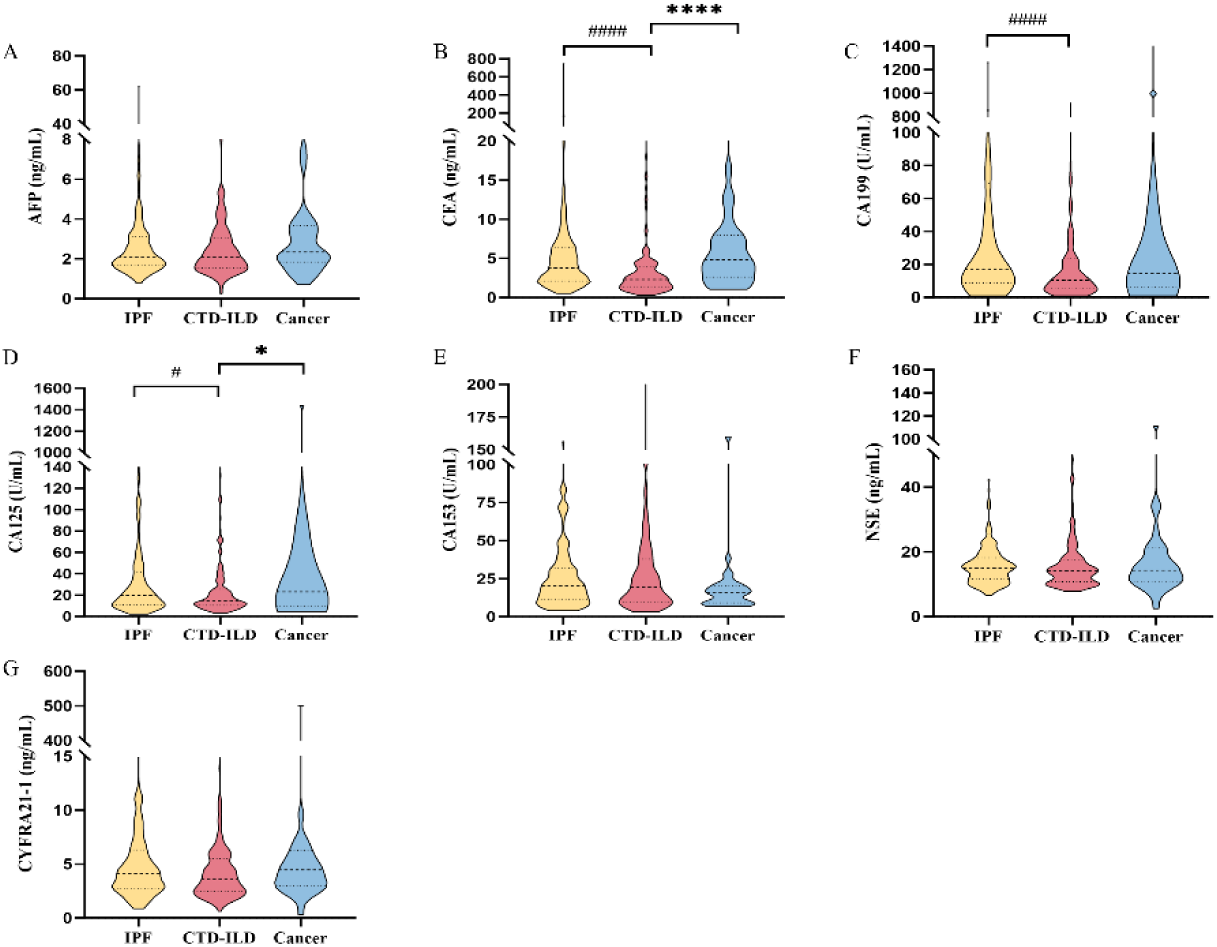
Comparison of Tumor Marker Levels Among Groups.(A)AFP, (B)CEA, (C)CA199, (D)CA125, (E)CA153, (F)NSE, (G)CYFRA21-1. The dotted line indicates the median. The upper and lower lines indicate the interquartile range. * indicates *p*<0.05 for Cancer-ILD vs. CTD-ILD comparison, **** indicates *p*<0.0001 for Cancer-ILD vs. CTD-ILD comparison, ^#^ indicates *p*<0.05 for IPF vs. CTD-ILD comparison, ^####^ indicates *p*<0.0001 for IPF vs. CTD-ILD comparison.

### 3.2. Analysis of tumor markers between CTD-ILD subgroups

CTD-ILD patients were classified into six clinical subtypes. Kruskal–Wallis tests showed no significant differences in AFP, CEA, CA199, CA125, CA153, or CYFRA21-1 among subtypes. NSE showed a nominal difference (χ² = 11.6, df = 5, unadjusted p = 0.0409), but this was not significant after Benjamini–Hochberg correction. Detailed values are presented in Table 2.

**Table 1.**
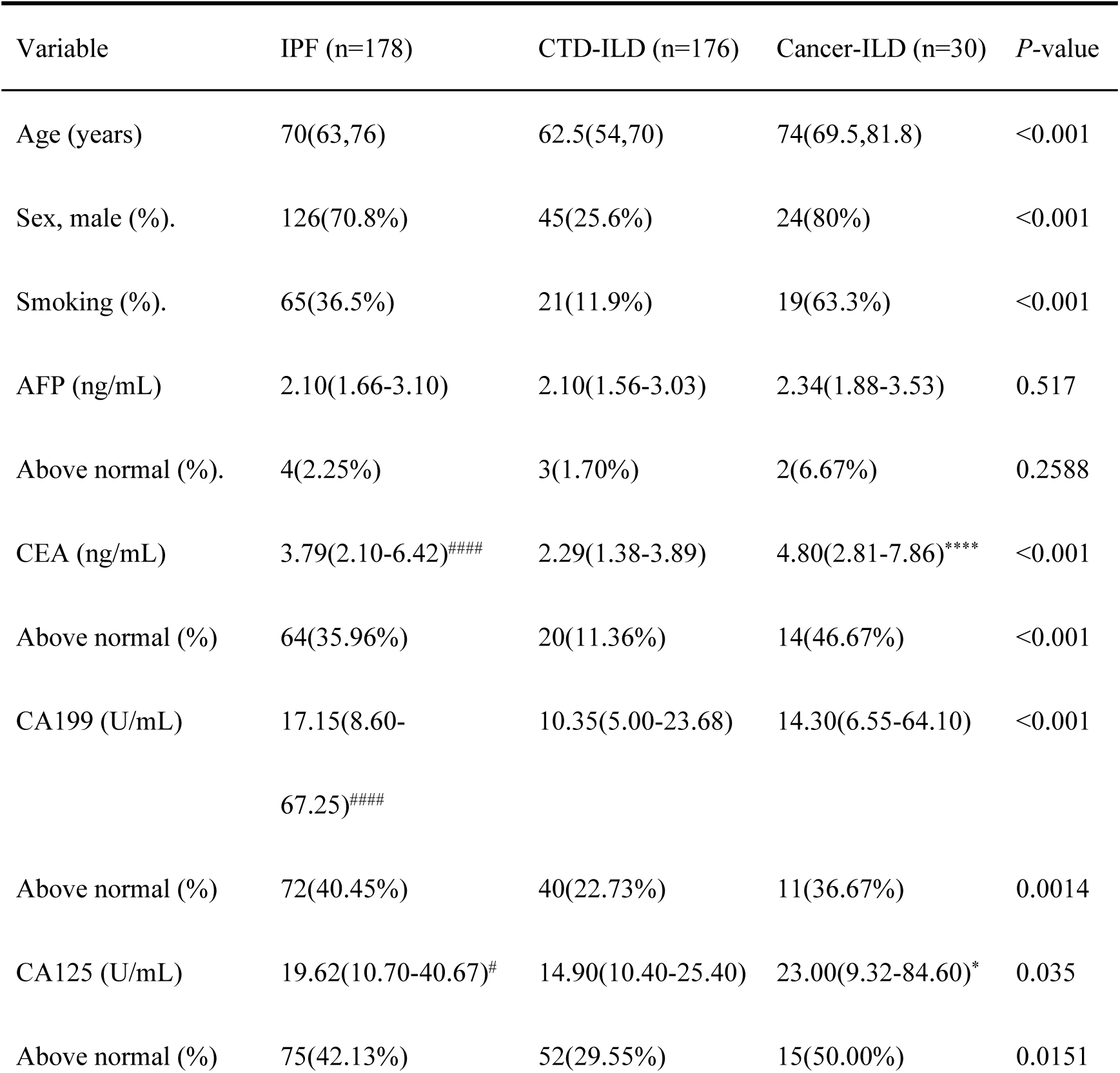
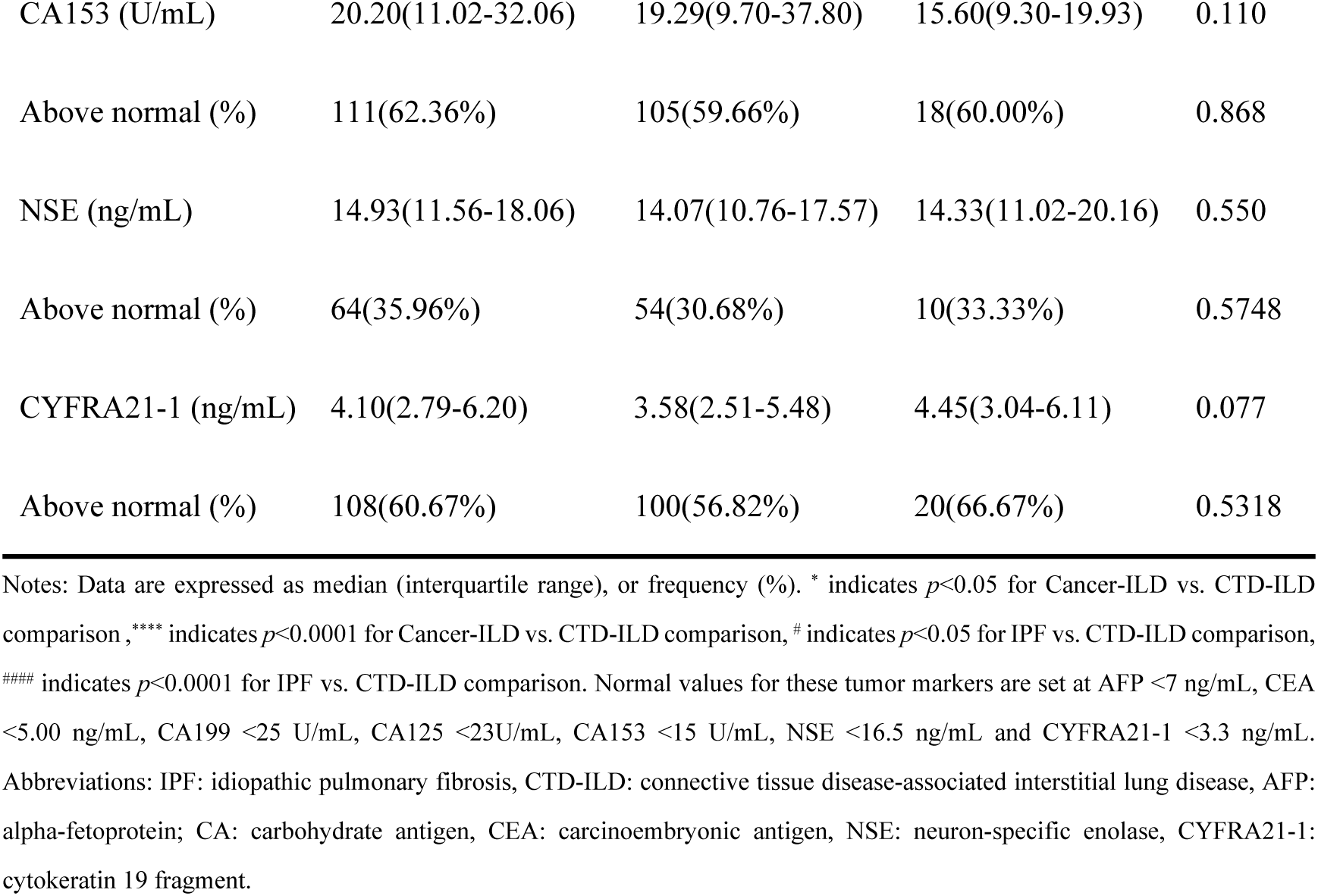
Comparison of baseline characteristics and tumor markers between the three groups.

**Table 2.**
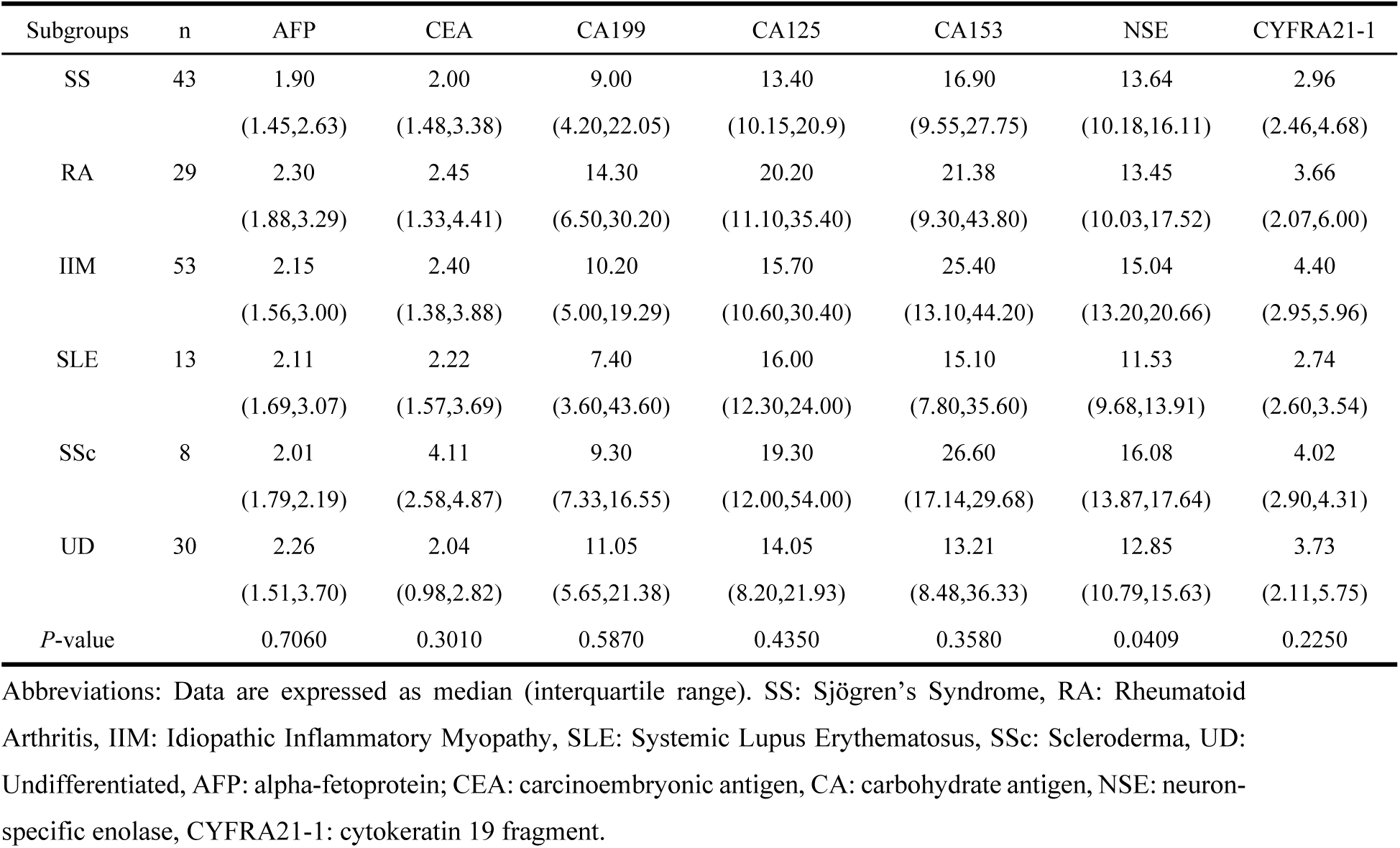
Comparison of tumor markers between CTD-ILD subgroups.

### 3.3. Correlation of Tumor Markers with Gas Exchange and Inflammatory Parameters in IPF and CTD-ILD

In IPF, CEA correlated negatively with PaO_2_ (r = −0.156, p = 0.038), and NSE also negatively correlated with PaO_2_ (r = −0.220, p = 0.003). No significant correlations were observed with PaCO_2_ or ESR. In CTD-ILD, NSE correlated negatively with PaCO_2_ (r = −0.181, p = 0.016) and CYFRA21-1 correlated negatively with PaO_2_ (r = −0.200, p = 0.008), while no significant correlations with ESR were found (Table 3).

**Table 3.**
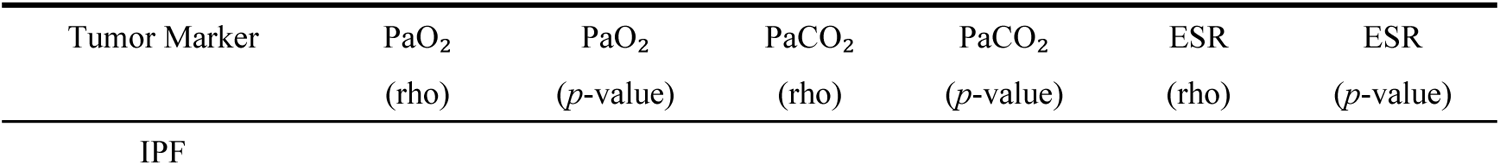
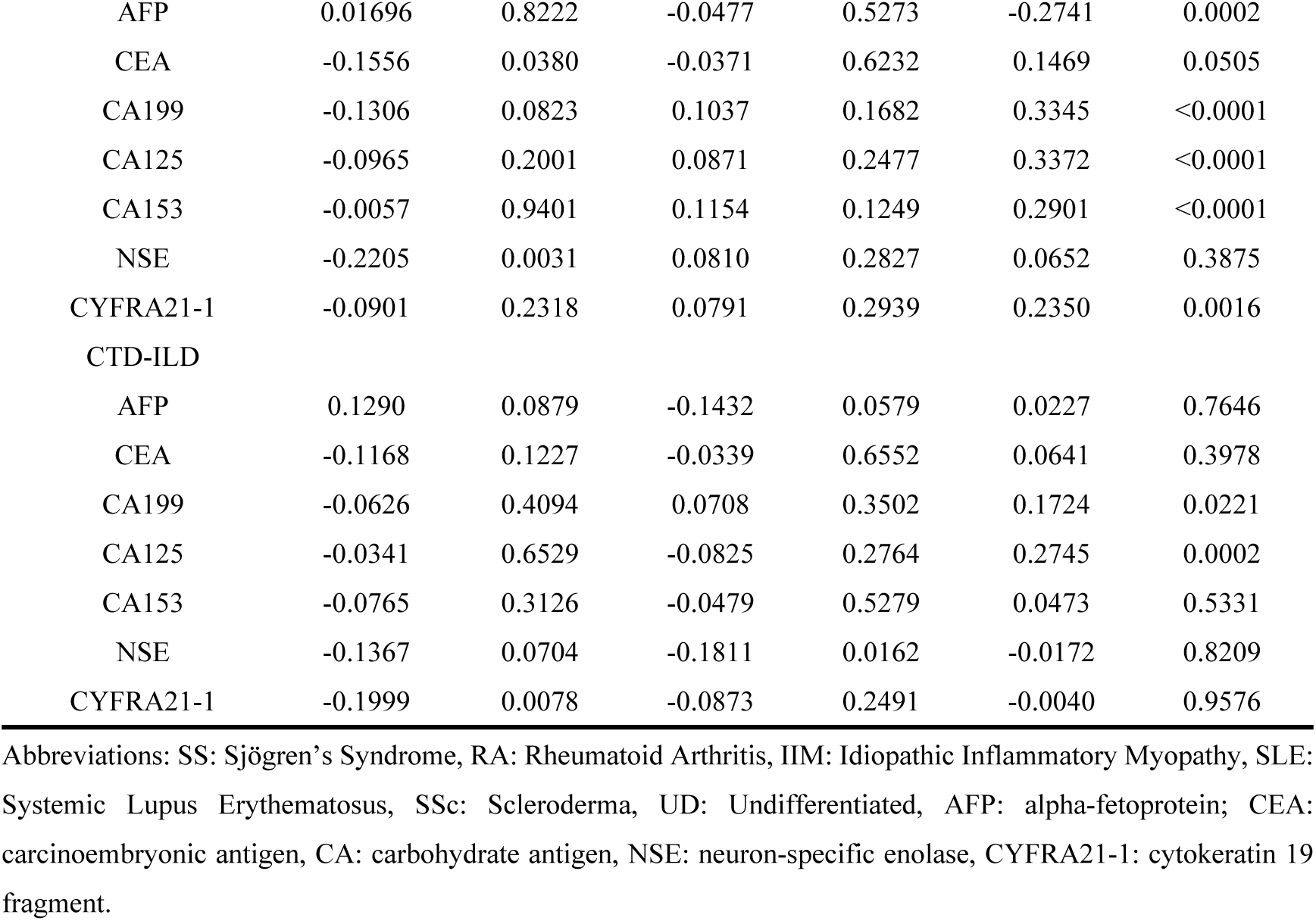
Correlation Analysis Between Tumor Markers, PaO_2_, PaCO_2_, and ESR in IPF and CTD-ILD Groups.

### 3.4. Clinical Parameters and Tumor Markers for Differential Diagnosis of IPF and CTD-ILD

Univariate logistic regression identified age, sex, smoking history, PaO_2_, CEA, and CA199 as significantly associated with IPF (p < 0.05). Other variables were not significant (Table 4).

**Table 4.** Logistic Regression Analysis Results for Clinical Parameters and Tumor Markers in IPF and CTD-ILD Groups.

| Variable | OR | 95%CI | P-value |
| --- | --- | --- | --- |
| sex(male) | 7.054 | 4.452-11.365 | <0.0001 |
| age | 1.068 | 1.046-1.092 | <0.0001 |
| Smoking(Yes) | 4.246 | 2.490-7.490 | <0.0001 |
| PaO<sub>2</sub> | 0.991 | 0.983-0.999 | 0.0245 |
| PaCO<sub>2</sub> | 0.997 | 0.969-1.026 | 0.8358 |
| ESR | 0.999 | 0.992-1.006 | 0.7865 |
| AFP | 1.068 | 0.989-1.214 | 0.2194 |
| CEA | 1.117 | 1.048-1.202 | 0.0015 |
| CA199 | 1.003 | 1.001-1.005 | 0.0159 |
| CA125 | 1.004 | 0.999-1.010 | 0.1594 |
| CA153 | 0.996 | 0.988-1.004 | 0.3520 |
| NSE | 0.989 | 0.963-1.014 | 0.3975 |
| CYFRA21-1 | 1.001 | 0.986-1.017 | 0.8680 |

Abbreviations: CI: Confidence Interval, AFP: alpha-fetoprotein; CEA: carcinoembryonic antigen, CA: carbohydrate antigen, NSE: neuron-specific enolase, CYFRA21-1: cytokeratin 19 fragment.

A multivariable diagnostic model was developed via binary logistic regression, including variables based on clinical relevance and univariate significance. Internal validation was performed using bootstrap resampling (1000 iterations) to assess calibration. The final model: Model1=−4.711+0.055×CEA+0.003×CA199+1.683×sex(male)+0.057×age+1.279×smoking(yes)−0.00 08×PaO2−0.0117×ESR.

ROC analysis showed an AUC of 0.833 (95% CI 0.770–0.876) with sensitivity 73.6% and specificity 82.4% at a cutoff of 0.467 (Table 5, Figure 3). No multicollinearity was observed (VIF < 2, tolerance > 0.1). Hosmer–Lemeshow test indicated acceptable fit (p = 0.068), and calibration intercept and slope were 0.0150 and 0.9702, respectively, indicating good agreement between predicted and observed probabilities.

**Figure 3.**
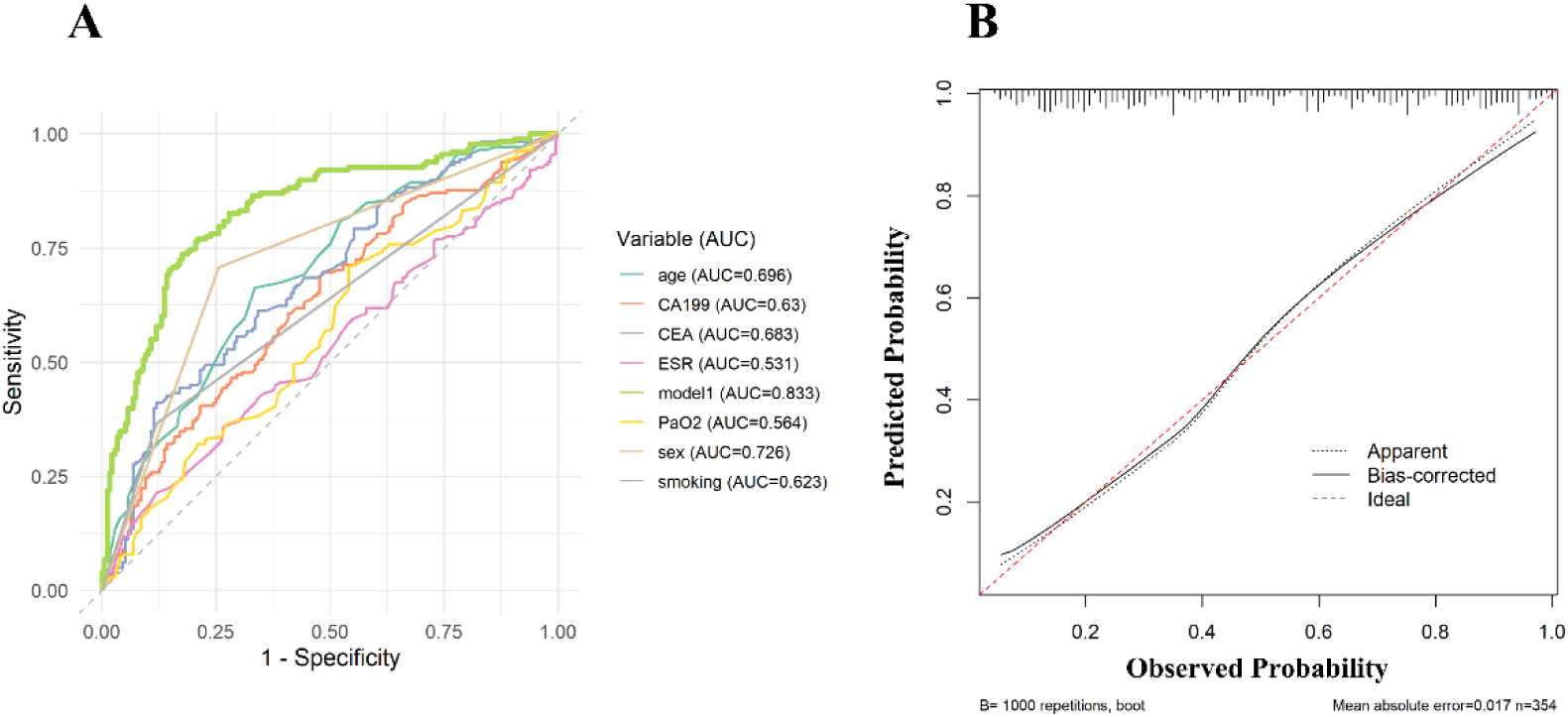
Diagnostic performance of multivariate model (A) ROC curve comparison of Model 1 with CEA 、 CA199、sex、age、smoking、PaO2、ESR for distinguishing IPF from CTD-ILD. (B) Calibration plot of the multivariable Model 1.

**Table 5.**
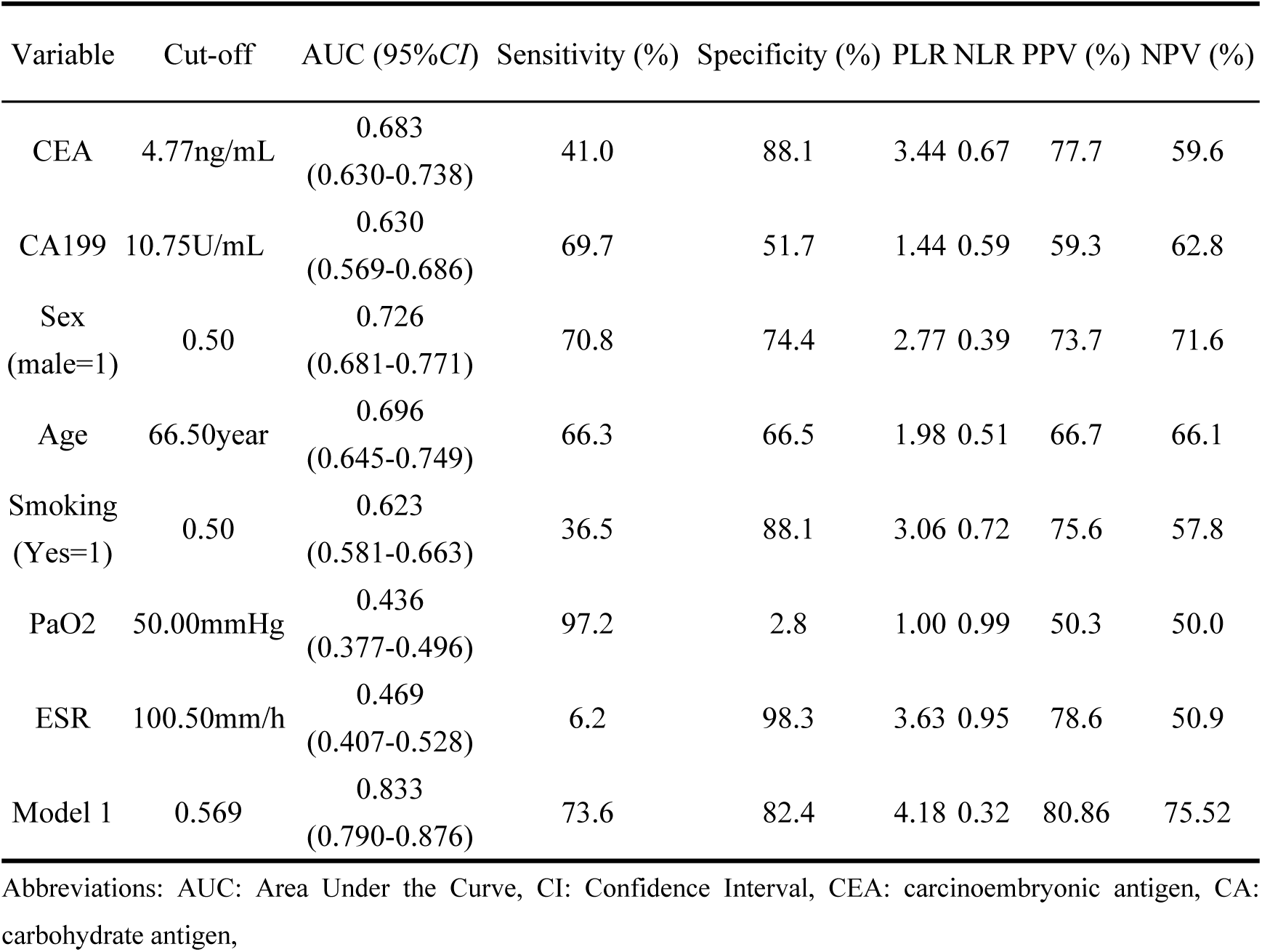
ROC Curve Analysis Results for Clinical Parameters and Tumor Markers in IPF and CTD-ILD Groups.

| Variable | Cut-off | AUC (95%CI) | Sensitivity (%) | Specificity (%) | PLR | NLR | PPV (%) | NPV (%) |
| --- | --- | --- | --- | --- | --- | --- | --- | --- |
| CEA | 4.77ng/mL | 0.683<br>(0.630-0.738) | 41.0 | 88.1 | 3.44 | 0.67 | 77.7 | 59.6 |
| CA199 | 10.75U/mL | 0.630<br>(0.569-0.686) | 69.7 | 51.7 | 1.44 | 0.59 | 59.3 | 62.8 |
| Sex<br>(male=1) | 0.50 | 0.726<br>(0.681-0.771) | 70.8 | 74.4 | 2.77 | 0.39 | 73.7 | 71.6 |
| Age | 66.50year | 0.696<br>(0.645-0.749) | 66.3 | 66.5 | 1.98 | 0.51 | 66.7 | 66.1 |
| Smoking<br>(Yes=1) | 0.50 | 0.623<br>(0.581-0.663) | 36.5 | 88.1 | 3.06 | 0.72 | 75.6 | 57.8 |
| PaO<sub>2</sub> | 50.00mmHg | 0.436<br>(0.377-0.496) | 97.2 | 2.8 | 1.00 | 0.99 | 50.3 | 50.0 |
| ESR | 100.50mm/h | 0.469<br>(0.407-0.528) | 6.2 | 98.3 | 3.63 | 0.95 | 78.6 | 50.9 |
| Model 1 | 0.569 | 0.833<br>(0.790-0.876) | 73.6 | 82.4 | 4.18 | 0.32 | 80.86 | 75.52 |

Abbreviations: AUC: Area Under the Curve, CI: Confidence Interval, CEA: carcinoembryonic antigen, CA: carbohydrate antigen,

## 4. Discussion

In this study, we found that some serum tumor markers were elevated in patients with ILD. Among them, CEA, CA199, and CA125 levels were significantly higher in patients with IPF than in those with CTD-ILD. Furthermore, the multivariable model incorporating tumor markers and clinical parameters demonstrated good diagnostic performance in distinguishing IPF from CTD-ILD. These findings suggest that readily available tumor markers may serve as useful adjuncts for differential diagnosis.

Previous studies have shown that the levels of tumor markers in patients with ILD are greater than those in healthy people^[11, 13]^. He et al. suggested that the serum levels of CA125, CA153 and CA199 could be used as relevant markers for SLE-ILD^[14]^ Bao et al. reported that higher levels of CA153 and CYFRA21-1 indicate an increased risk of ILD in CTD patients^[13]^. Wei et al. reported that the levels of tumor markers (e.g., CEA, CA153, and CA125) were greater in patients with primary Sjögren’s syndrome complicated with interstitial lung disease than in non-ILD patients^[12]^. This is similar to what we found in this study. In this study, we found that tumor marker levels in patients with ILD were frequently elevated above the normal range and that the serum levels of CEA, CA199, and CA125 were significantly greater in patients with IPF than in those with CTD-ILD.

The mechanisms underlying the elevation of tumor markers in ILD remain incompletely understood; however, several pathophysiological processes may contribute. ILD is characterized by recurrent alveolar epithelial injury and aberrant repair, leading to chronic inflammation and fibrogenesis^[22]^. This persistent inflammatory milieu promotes macrophage activation and the release of cytokines such as IL-6, IL-1, IL-10, and TNF-α, which in turn stimulate neutrophil infiltration and the abnormal proliferation and differentiation of alveolar epithelial cells and fibroblasts, thereby enhancing the production and release of tumor markers^[23]^. In IPF specifically, repeated alveolar epithelial injury and dysregulated repair represent the central pathogenic axis. Damage to type II alveolar cells and bronchiolar epithelium can result in the release of epithelial-derived antigens such as CEA and CA199, while sustained EMT and fibroblast activation further perpetuate extracellular matrix deposition and fibrosis^[24, 25]^. CA125 (MUC16), a high-molecular-weight glycoprotein expressed on mesothelial and epithelial surfaces, is similarly upregulated in response to serosal inflammation and fibrotic remodeling^[26][27]^. In contrast, CTD-ILD pathogenesis is primarily driven by lymphocytic infiltration and immune complex deposition, with epithelial injury playing a secondary role^[28]^ This fundamental mechanistic difference likely explains the comparatively lower levels of epithelial-derived tumor markers (CEA and CA199) observed in CTD-ILD patients compared with those with IPF.

To account for the confounding influence of malignancy on tumor marker levels, we included a cancer-ILD group in our analysis. In clinical settings, many ILD patients may have concurrent cancers, which can significantly increase the levels of markers such as CEA and CA199. By comparing the three groups, we found that CEA, CA199 and CA125 levels in IPF patients were significantly higher than those in CTD-ILD patients but lower than those in cancer-ILD patients. The observed gradient in tumor marker levels supports the conclusion that IPF itself contributes to the elevation of these markers through specific pathophysiological mechanisms. The fact that IPF levels are lower than those in cancer-ILD patients helps rule out malignancy as the primary cause and suggests the involvement of IPF-specific factors.

To further differentiate interstitial lung diseases of distinct etiologies, we compared tumor marker levels across clinical subtypes within the CTD-ILD group. A detailed subgroup analysis was conducted on the basis of six defined CTD-ILD subtypes. Despite the nonnormal distributions of tumor markers, no significant differences were found in the levels of AFP, CEA, CA199, CA125, CA153, or CYFRA21-1 across the subtypes, suggesting that these markers may not be sensitive enough to capture the heterogeneity of CTD-ILD. This finding is in line with those of previous studies indicating that the utility of tumor markers in patients with CTD-ILD may be constrained by the complexity and varied pathophysiology of these diseases. However, it is worth noting that NSE levels significantly varied across subtypes, particularly in the myositis group. Among the CTD-ILD subtypes, the myositis subgroup presented higher NSE levels than the other subgroups did, as shown by unadjusted analysis. This result is consistent with the known role of NSE as a marker of neuronal and muscular injury and supports the hypothesis that myositis-related ILD may involve increased muscle damage, which could contribute to the observed elevation in NSE levels^[29]^.

Our research investigated the relationships between tumor markers and blood gas parameters (PaO_2_, PaCO_2_) as well as the ESR. In the IPF group, CEA was significantly negatively correlated with PaO_2_, indicating that the higher the CEA level was, the more severe the degree of oxygen damage was. These findings support the role of CEA as a potential marker of disease severity in IPF. Similarly, NSE was negatively correlated with PaO_2_, which was consistent with the correlations between disease progression and reduced lung function. In the CTD-ILD group, NSE was negatively correlated with PaCO_2_, suggesting that higher NSE levels might be associated with impaired alveolar ventilation and carbon dioxide removal. CYFRA21-1 is negatively correlated with PaO_2_. Since CYFRA21-1 indicates epithelial cell renewal, this may suggest that greater lung epithelial damage or remodeling is associated with the deterioration of hypoxemia. This highlights its potential as a marker of disease severity and gas exchange problems in patients with CTD-ILD.^[30]^ Overall, these correlations suggest that tumor markers can be used to assess the severity of ILD and respiratory function.

In this study, we evaluated the value of various serum biomarkers and clinical factors in differentiating IPF from CTD-ILD. Univariate logistic regression identified several potential predictive factors, including CEA, CA199, PaO_2_, age, ESR, sex and smoking history. Among these, sex, age, CEA, and CA199 exhibited moderate diagnostic performance, with AUC values of 0.762 (95% *CI*, 0.681–0.771), 0.696 (95% *CI*, 0.645–0.749), 0.683 (95% *CI*, 0.630–0.738) and 0.630 (95% *CI*, 0.569–0.686), respectively. Although the ESR was not statistically significant in the univariate analysis, we incorporated it into our multivariate model because of its well-established clinical relevance in patients with CTD-ILD. To improve diagnostic discrimination between IPF and CTD-ILD, we developed a multivariate logistic regression model (Model 1). This integrated model demonstrated robust discriminative capacity, achieving an AUC of 0.833 (95% CI: 0.790–0.876) with 73.3% sensitivity and 82.4% specificity, significantly outperforming all individual predictors. Validation analysis confirmed its consistent performance in differentiating these disease entities.

There are several limitations to this study. First, it is a single-center, retrospective analysis with a modest sample size, which may limit external generalizability and increase the risk of selection bias. Second, the retrospective design precludes causal inference and may introduce information bias from incomplete or inconsistent medical records. Third, we did not include conventional immunological indicators such as ANA, ENA, MSA, RF, or CCP, nor did it analyze detailed clinical features, which may provide additional diagnostic value in differentiating CTD-ILD from IPF. Future studies incorporating serological, clinical, and imaging parameters are warranted to further validate and expand upon our findings. Fourth, tumor marker measurements were captured at a single time point on admission and longitudinal kinetics were not assessed; temporal trends might be informative for disease activity or prognosis. Fifth, heterogeneity in laboratory assays, potential preanalytical variability, and unmeasured comorbidities (including occult malignancy or inflammatory conditions) could confound marker levels.

To mitigate these limitations in future studies, we plan prospective, multicenter validation with standardized assay platforms, serial sampling to evaluate marker dynamics, inclusion of comprehensive autoantibody panels and structured clinical phenotyping, and pre-specified adjustment for key confounders. Such studies will clarify whether tumor markers provide independent or complementary diagnostic value—particularly in seronegative or clinically ambiguous ILD cases.

## 5. Conclusion

This study revealed significantly higher serum levels of CEA, CA199, and CA125 in patients with IPF than in those with CTD-ILD. CEA and CA199 served as independent predictors for differentiating the two diseases, and their combination with clinical indicators further improved diagnostic performance. In IPF, CEA and NSE levels were negatively correlated with PaO_2_, whereas in CTD-ILD, NSE was negatively correlated with PaCO_2_ and CYFRA21-1 with PaO_2_, suggesting their potential relevance to gas-exchange impairment and disease severity. These findings indicate that serum tumor markers may serve as valuable complementary tools for the diagnosis and stratification of ILD subtypes. Validation in large-scale, prospective, multicenter studies is warranted.

## Credit authorship contribution statement

**Yingying Du**: Conceptualization, Validation, Formal analysis, Data curation, Writing—original draft. **Xuxiang Song**: Methodology, Validation. **Qunli Ding**: Resources, supervision, conceptualization, writing-review & editing.

## Data Availability

The data underlying this study cannot be shared publicly due to ethical and legal restrictions related to patient privacy. However, the data are available from the the First Hospital Affiliated with the Ningbo University Ethics Committee (contact via) for researchers who meet the criteria for access to confidential data.

## Acknowledgments

This work was supported by the Natural Science Foundation of Ningbo (No. 2023J156) and the Key Technology of KeChuang Yongjiang 2035 (No. 2024Z182).

## Ethics approval

The study followed the principles set forth in the Declaration of Helsinki and was approved by the Institutional Review Board of the First Hospital Affiliated with Ningbo University. (Approval No.2025 Research No. 129RS). The need for informed consent was waived by the Ethics Committee because of the retrospective design of the study.

## Consent to participate

Informed consent was obtained from all individual participants included in the study.

